# Anti-Hepatitis C Antibody Carriage and Risk of Liver Impairment in Rural-Cameroon: Adapting the Control of Hepatocellular Carcinoma for Resource-Limited Settings

**DOI:** 10.1101/2023.08.16.23294167

**Authors:** Rodrigue Kamga Wouambo, Gaelle Panka Tchinda, Luc Aime Kagoue Simeni, Paule Dana Djouela Djoulako, Clarisse Irene Yateu Wouambo, Ghislaine Flore Tamko Mella, Eric pascal Tchoumi Leuwat, Djoda Bello, Joseph Fokam

## Abstract

**Background:** The global Viral hepatitis elimination by 2030 is uncertain in resource-limited settings (RLS), due to high burdens and poor diagnostic coverage. This sounds more challenging for hepatitis C virus (HCV) given that antibody (HCVAb) sero-positivity still lacks wide access to HCV RNA molecular testing. This warrants context-specific strategies for appropriate management of liver impairment in RLS. We herein determine the association between anti-HCV positivity and liver impairment in an African RLS.

**Methods:** A facility-based observational study was conducted from July-August 2021 among individuals attending the “St Monique” Health Center at Ottou, a rural community of Yaounde,Cameroon. Following a consecutive sampling, consenting individuals were tested for anti-HCV antibodies, hepatitis B surface antigen (HBsAg) and HIV antibodies (HIVAb) as per the national guidelines. After excluding positive cases for HBsAg and/or HIVAb, liver function tests (ALT/AST) were performed on eligible participants (HBsAg and HIVAb negative) and outcomes were compared according to HCVAb status; with p<0.05 considered statistically significant.

**Results:** Out of 306 eligible participants (negative for HBsAg and HIVAb) enrolled, the mean age was 34.35±3.67 years. 252(82.35%) were female and 129 (42.17%) were single. The overall HCVAb sero-positivity was 15.68%(48/306), with 17.86% (45/252) among women vs. 5.55%(3/54) among men [OR (95%CI)=3.69(2.11-9.29),*p*=0.04]. HCVAb Carriage was greater among participants aged >50 years compared to younger ones [38.46%(15/39) versus 12.36% (33/267) respectively, OR(95%CI)=4.43(2.11-9.29), p<0.000] and in multipartnership [26.67%(12/45)vs.13.79%(36/261) monopartnership, OR (95%CI)= 2.27(1.07-4.80),*p*=0.03]. The liver impairment rate (abnormal ALT+AST levels) was 30.39%(93/306), with 40.19%(123/306) of abnormal ALT alone. Moreover, the burden of Liver impairment was significantly with aged>50 versus younger ones [69.23% (27/39) versus 24.72%(66/267) respectively, p<0.000). Interestingly, the burden of liver impairment (abnormal AST+ALAT) was significantly higher in HCVAb positive (62.5%, 30/48) versus HCVAb negative (24.42%, 63/258) participants, OR: 3.90 [1.96; 7.79], p=0.0001.

**Conclusions:** In this rural health facility, HCVAb is highly endemic and the burden of liver impairment is concerning. Interestingly, HCVAb carriage is associated with abnormal liver levels of enzyme (ALT/AST), especially among the elderly populations. Hence, in the absence of nuclei acid testing, ALT/AST are relevant sentinel markers to screen HCVAb carriers who require monitoring/care for HCV-associated hepatocellular carcinoma in RLS.

## BACKGROUND

Hepatitis C is a liver inflammation caused by a hepatitis C virus (HCV). This virus causes acute hepatitis that evolves in majority (85%) into chronic hepatitis [1]. Chronic hepatitis C is one of the main causes of cirrhosis and primary liver cancer. Globally, the World Health Organization (WHO) estimates that an estimated 58 million people have chronic hepatitis C virus infection, with about 1.5 million new infections occurring per year [2]. In 2019, approximately 290,000 people died from hepatitis C, mostly from cirrhosis and hepatocellular carcinoma [3]. In 2016, World Health Assembly has adopted the Global Health Sector Strategy (GHSS) on viral hepatitis to eliminate hepatitis by 2030 [4] However, available evidence demonstrates that global viral hepatitis elimination by 2030 is highly unlikely especially in Low and middle income countries where rates of hepatitis B and C diagnosis are very low, averaging 8% and 18%, respectively[5]. In fact, for HCV infection, 80% of high-income countries are not on track to meet HCV elimination targets by 2030, and 67% will not meet elimination targets even if they were given an additional 20 years [6]. In contrast to HBV, there is no prophylactic HCV vaccine [7].

In Cameroun, hepatitis C is endemic and the prevalence of HCV varies widely between 1 and 23.9% depending on the study population [8, 9, 10, 11]. WHO and The Ministry of Public Health of Cameroon have made the fight against viral hepatitis their focus through vast treatment programs for hepatitis B/C and that have now been progressively made accessible to all social layer at low cost [12]. However, the lack of national Hepatitis C screening and treatment guidelines is reflected by a diversity in diagnostic and treatment protocols across care providers, resulting in unnecessary costs and possible sub-optimal clinical outcomes. Additionally, central organisation of Hepatitis C care, insufficient technical and human capacity for HCV testing, alongside high costs associated with diagnosis and treatment (generally paid out-of-pocket), substantially limit access to HCV diagnosis and care [13]. Most PLHCV in Cameroon are unaware of their status, with a negative impact on transmission and disease progression [14-16].

In community areas, in addition to the to lack of awareness on HCV, the problem is compound by low quality of health care, poverty and underdevelopment that constitute significant obstacles to early diagnosis and adequate management of HCV cases. In those settings, many HVCAb positive patients by rapid diagnostic test are either orientated for enrolment to a distant reference hospital center or requested an expensive and time-consuming HCV RT-PCR confirmatory test from a reference laboratory. This situation increases loss to follow-up of infectious patients with sustained HBV viral load who generally get back to health facilities afterwards with complications such as cirrhosis or hepatocellular cancer. Indeed, elevated HCV viral load is described to be consistently associated with high infectivity and risk of HCC occurrence [17-19]. Thus, much efforts should foster early identification and management of infected cases. Alanine Amino transferase (ALT) is a valuable liver enzyme test to detect otherwise inapparent liver disease [20]. Its elevation is considered as an indicator of Liver Damage [20, 21] and could also predict viremia in anti-HCV-positive patients [22]. Besides, ALT measurement affords a readily available, low-cost blood test that is utilized throughout many countries as a tool for detection of liver disease [22].

We conducted a mass screening of HCV in Ottou village-Yaounde that aimed at determining the association between HCVAb positivity and liver impairment in an African RLS.

## MATERIALS AND METHODS

### A. Study design and population

A cross-sectional study was conducted during a health campaign held from the 12 July to 12 August 2021 at the “Sainte Monique-Ngonmeda” Health Center, located at Ottou-village, at the periphery of Yaounde Cameroon. Ottou is a small village not so far from Yaounde and has been choosed for its hererogeneous population and for its geographical position with the presence of health center and a private medical laboratory capable to process ALT/AST biochemical test.

After getting required administrative authorization, consenting inhabitants of Ottou aged >18years, were enrolled consecutively at “Sainte Monique-Ngonmeda” Health Center for the health campaign. Each participant always had the choice either to participate only to the campaign or to both, the campaign and the study. Those who signed the informed consent, filled out the structured questionnaire in presence of a member of the team were included in the study. Conversely, those who declined the invitation and/or refused to sign the informed consent form could participate to the health campaign but were excluded from the study. From each participant included in the study, blood was collected onsite by a prick on the middle finger for a rapid detection of HCV antibodies, HIV antibodies and HBsAg. In parallel, 5ml whole blood was also collected and stored in a cooler at+4 °C before being transferred to the private laboratory “MEDIBIO-LAB” for ALT/AST dosage.

### B. Ethics Approval and Consent

The study was conducted in accordance with the Declaration of Helsinki. The research proposal was evaluated and ethical approval was obtained from the Regional Ethics Committee for Research involving humans of the Center Region Cameroon (N°/CE01132/CRERSHC, on May 10^th^, 2021). Additionally, we obtained administrative authorization from the directors of “Sainte Monique” Health Center (Ref N°115/2021/CSGPSM/RS) and MEDIBIO-LAB (N°2021/10/MEDIBIO-LAB/LR) where the study was conducted. An information note was given to all the eligible participants, who then provided their written informed consent before enrollment into the study. The confidentiality of study participants was secured via the use of identification codes.

### C. Determination of Minimum Sample Size

The minimum sample size was obtained using the standard formula:

n= z^2^.p (1-p)/m^2^, with “z” = the standard deviation of 1.96 (95% confidence interval); “p”= Estimated nationwide seroprevalence of HCV in Cameroonians subjects aged >15 years reported by Bigna et al (6.5%) [8]; “m”=error rate of (5%) and “n”= minimum sample size.

### D. Sample Collection and Conservation

Eligible participants who gave their approval were subjected to rapid diagnostic testing and blood collection for further testing (ALT/AST).

#### 1. Blood Sampling and rapid testing “on-site”

Blood was collected in aseptic conditions. In all, One drop of whole blood was required for the “on-site” testing for each rapid detection of HCV antibodies (Cypress diagnostics Technologies Inc…, USA) HIV antibodies (Determine HIV 1/2; Alere Medical Co., Chiba, Japan) and HBsAg (DiaSpot HBsAg; DiaSpot Diagnostics, USA) according to the SOP /presented to us by the manufacturer. Cypress Diagnostics® HCV Rapid Antibody Test is rapid diagnostic test for the qualitative detection of HCV antibodies in serum and whole blood. Independent evaluation reported 99.4% sensitivity of this test in fingerstick and 97.6% sensitivity in whole blood [24].

#### 2. Samples Transportation

Antibodies testing was performed on-site, whereas whole blood collected in dry tubes were transported to MEDIBIO-LAB, located not so far, for liver enzymes (ALT/AST) dosage. All blood samples were temporarily stored between 2 and 8 °C immediately after collection. And then, directly transferred to MEDIBIO-LAB and analyzed upon arrival.

#### 3. Transaminases Testing

Transminases (ALT, AST) were analysed from blood samples of HBsAg and HIVAb negative participants as described previously [25]. In Brief, after centrifugation 2000rpm/min during 5min, transaminases level were tested in each serum samples using an autoanalyzer. The level of ALT >33 U/L and AST >31 U/L for females and ALT >40 U/L and AST >37 U/L for males were classified as abnormal. Laboratory quality control will be done regularly to verify inter- and intra-assay reproducibility.

#### 4. Data analysis

The analyses were performed using the software package Stat view 5.1. The continuous variables are presented in terms of mean ± Standard deviation (Std) and categorical variables in absolute number (proportion in %). Given that data on some variables were not properly recorded for all patients, we decided to include only participants with all information in the analysis. For all the analyses, the significance level was set at p<0.05.

#### 5. Limitation

The main limitation of this study was the absence of HCV RNA molecular testing

## Results

### 1. Baseline characteristics of the study population

The table 1 below presents the sociodemographic characteristics of the study population

**Table 1:** Sociodemographic Characteristics of the Study population.

|  |  | Number | Percentage (%) |
| --- | --- | --- | --- |
| Sex | Female | 252 | 82.35 |
|  | Male | 54 | 17.65 |
| Age | <30 | 156 | 50.98 |
|  | [30-50[ | 111 | 36.27 |
|  | >50 | 39 | 12.74 |
| Matrimonial status | Single | 129 | 42.16 |
|  | Married | 168 | 54.90 |
|  | Widow(er) | 09 | 2.94 |
| Knowledge about the disease | Yes | 255 | 83.33 |
|  | No | 51 | 16.67 |
| history of blood transfusion | Yes | 12 | 3.92 |
|  | No | 294 | 96.08 |
| Regular use of condoms | Yes | 21 | 6.86 |
|  | No | 285 | 93.14 |
| **HCV history in the neighbourhood** | Yes | 63 | 20.58 |
|  | No | 243 | 79.41 |
| **Number of sex partners** | Mono-partnership | 261 | 85.29 |
|  | Multi-partnership | 45 | 14.71 |
| **Sterilisation of hairdressing materials** | Yes | 159 | 51.96 |
|  | No | 147 | 48.04 |
| **Alcohol consumption** | Yes | 228 | 74.51 |
|  | No | 78 | 25.49 |
| **History of sexually transmitted infections** | Yes | 129 | 42.16 |
|  | No | 177 | 57.84 |
| **Clinical signs of hepatitis C** | Yes | 294 | 90.20 |
|  | No | 30 | 9.80 |
| **Piercing** | Yes | 09 | 2.94 |
|  | No | 297 | 97.06 |
| **Illicit Drug consumption** | Yes | 00 | 00 |
|  | No | 306 | 100 |

A total of 306 participants were surveyed between 12 July and 12 August 2021 at the “Sainte Monique-Ngonmeda” Health Center, Ottou-village, at the periphery of Yaounde Cameroon. In this study, the female participants were predominant with a percentage of 82.35% (n=252), versus 17.65% (n=54) for males participants with a sex ratio (F/M) of 5/1. The mean age was 34.52±3.42 years [min 18, max 72] and subjects aged under 30 years were the most represented. Moreover, more than half (54.9%) of the participants were married, and 42.16 % were single and 2.94% widowed. The majority of Participants declared to have the knowledge on the disease (83.33%) and only 3.92% had a history of blood transfusion. Regular use of condoms, HCV history in the neighborhood, multi-partnership, history of sexually transmitted infections and piercing were recorded in 6.86%, 20.59%, 14.71%, 42.16% and 2.94% of the participants respectively. Lastly, 74.50% were alcohol consumers and none was illicit drug users. In this study, 90.20% of the participants had clinical signs of hepatitis C (see Table 1).

### 2. Prevalence of HCVAb and risk factors

This table below summarized the prevalence of HCVAb according to risk factors

According to the hepatitis C antibody rapid detection tests results, participants who tested positive were 48 of the 306, for a prevalence 15.69% [11.58% -19.78%]. Univariate analysis shows a higher seroprevalence of HCVAb in female subjects [17.86% vs. 5.55% men, OR (95%CI)= 3.69(2.11-9.29), *p*=0.04], subjects aged >50 years [OR(95%CI)=4.43(2.11-9.29), *p*<0.001], multipartnership [26.67% vs. 13.79% monopartnership, OR (95%CI)= 2.27(1.07-4.80), *p*=0.03] and subjects who didn’t regularly sterilized their hairdressing materials [22.45% vs. 9.43%people sterilising theirs, OR (95%CI)= 2.78 (1.44-5.36), *p*=0.003] (see Table 2).

**Table 2.** HCVAb carriage according to sociodemographic factors, behaviour and practices.

| **Variables** |  | **N=306** | **HCVAb**<br>**n(%)** | **OR** | **95%CI** | **p-value** |
| --- | --- | --- | --- | --- | --- | --- |
| **Sex** | Female | 252 | 45 (17.86) | 3.69 | [1.10-12.37] | **0.04** |
|  | Male | 54 | 3 (5.55) | Ref |  |  |
| **Age** | <30 | 156 | 24 (15.38) | Ref |  | **0.00** |
|  | [30-50[ | 111 | 9 (8.11) | 0.48 | [0.22- 1.09] |  |
|  | >50 | 39 | 15 (38.5) | 3.43 | [1.58-7.48] |  |
| **Matrimonial status** | Single | 129 | 24 (18.60) | 1.78 | [0.93-3.53] | 0.12 |
|  | Married | 168 | 21 (12.25) | Ref |  |  |
|  | Widow(er) | 09 | 3 (33.33) | 2.33 | [0.58-3.31] |  |
| **Knowledge about the disease** | Yes | 255 | 39 (15.29) | Ref | [0.38-1.82] | 0.84 |
|  | No | 51 | 9 (17.65) | 1.19 | [0.53-2.63] |  |
| **history of blood transfusion** | Yes | 12 | 3 (25.00) | 1.84 | [0.48-7.07] | 0.85 |
|  | No | 294 | 45(15.31) | Ref |  |  |
| **Regular use of condoms** | Yes | 21 | 3 (14.28) | 1.12 | [0.32-3.98] | 0.66 |
|  | No | 285 | 45 (15.79) | Ref |  |  |
| **HCV history in the neighborhood** | Yes | 63 | 09 (14.28) | 0.87 | [0.4-1.92] | 0.89 |
|  | No | 243 | 39 (16.05) | Ref |  |  |
| **Number of sex partners** | Mono-partnership | 261 | 36 (13.79) | 2.27 | [1.07-4.80] | 0.03 |
|  | Multi-partnership | 45 | 12 (26.67) | Ref |  |  |
| **Sterilisation of hairdressing materials** | Yes | 159 | 15 (9.43) | Ref |  |  |
|  | No | 147 | 33 (22.45) | 2.78 | [1.44-5.36] | 0.003 |
| **Alcohol consumption** | Yes | 228 | 39 (17.10) | 1.58 | [0.73-3.43] | 0.71 |
|  | No | 78 | 09 (11.54) | Ref |  |  |
| **History of sexually transmitted infections** | Yes | 129 | 24 (18.60) | 1.46 | [0.78-2.70] | 0.30 |
|  | No | 177 | 24 (13.56) | Ref |  |  |
| **Clinical signs of hepatitis C** | Yes | 297 | 48 (16.16) | 0.16 |  | 0.39 |
|  | No | 09 | 00 (00) | Ref |  |  |
| **Piercing** | Yes | 09 | 00 (00) | 00 |  | 0.39 |
|  | No | 297 | 48 (16.16) | Ref |  |  |
| **Illicit Drug consumption** | Yes | 0 | 00 | 00 |  | 0.94 |
|  | No | 306 | 48 (15.69) | Ref |  |  |

In multivariate analysis using logistic regression, only gender, age, multipartnership were found to be significantly associated to HCVAb carriage (p<0.000)

### 3. Liver Function of the study population

#### 3.1. Prevalence of liver damage

The table 3 shows the Prevalence of liver damage in the study population

**Table 3:** Prevalence of liver impairment.

| TRANSAMINASES | Normal | Abnormal |
| --- | --- | --- |
| N=306 | n(%) | n(%) |
| ALT | 183(59.80) | 123 (40.20) |
| AST | 162 (52.29) | 144 (47.06) |
| AST+ALT | 138 (45.09) | 93 (30.39) |

The prevalence of liver impairment (abnormal ALT+AST) was 30.39% (93/306) with 40.20% (123/306) of abnormal ALT (see table 3)

#### 3.2. Liver damage according to risk factors

The table 4 below shows liver impairment according to risk factors

**Table 4.** Liver damage according to risk factors.

|  |  |  | AST | ALT | ALT+AST |
| --- | --- | --- | --- | --- | --- |
|  |  |  | Abnormal | Abnormal | Abnormal |
|  |  |  | (N=144) | (N=123) | (N=93) |
|  |  |  | n(%) | n(%) | n(%) |
| Gender | F | 252 | 120 (47.62) | 101(40.08) | 73(28.97) |
|  | M | 54 | 24(44.44) | 22 (40.74) | 20(37.04) |
| p-value |  |  | 0.67 | 0.93 | 0.24 |
| Age | <30 | 156 | 55 (35.26) | 53 (33.97) | 33 (21.15) |
|  | [30-50[ | 111 | 57 (51.35) | 40 (70.17) | 33 (29.73) |
|  | >50 | 39 | 32 (82.05) | 30 (69.24) | 27 (69.23) |
| p-value |  |  | <0.001 | <0.001 | <0.001 |
| Matrimonial Status | Célibat | 129 | 51 (39.53) | 48(37.21) | 30(23.25) |
|  | Marié | 168 | 90 (53.57) | 72 (42.86) | 60(35.71) |
|  | Veuve | 09 | 3 (33.33) | 3 (33.33) | 3 (33.33) |
| p-value |  |  | 0.06 | 0.68 | 0.06 |
| HCV history in the neighborhood | Yes | 63 | 30(47.62) | 27(42.86) | 19(30.16) |
|  | No | 243 | 114(59.26) | 96(39.51) | 74(30.45) |
| p-value |  |  | 0.92 | 0.63 | 0.96 |
| **Alcohol consumption** | Yes | 228 | 115(50.44) | 100(43.86) | 75(32.89) |
|  | No | 78 | 29(37.18) | 23(29.49) | 18(23.08) |
| *p-value* |  |  | 0.04 | 0.03 | 0.1 |
| **History of sexually transmitted infections** | Yes | 129 | 62(48.06) | 48(37.21) | 35(27.13) |
|  | No | 177 | 82(46.33) | 75(42.37) | 58(32.77) |
| *p-value* |  |  | 0.76 | 0.36 | 0.29 |
| **Clinical signs of hepatitis C** | Yes | 297 | 139(46.80) | 120(40.4) | 90(30.30) |
|  | No | 09 | 5(55.56) | 3(33.33) | 3(33.33) |
| *p-value* |  |  | 0.86 | 0.93 | 0.86 |

It comes out from table 4 that liver damage (abnormal ALT+AST level) was associated to Age (p=0.01) with subject aged >50 most affected as compared to younger ones [69.23% (27/39) versus 24.72% (66/267) respectively, p<0.000).

#### 3.3. Liver impairment according to hepatitis C antibodies carriage (HCVAb)

This table presents liver impairment according to hepatitis C antibodies carriage (HCVAb)

The prevalence of liver damage (abnormal AST+ALAT) is much higher in HCVAb positive subjects (66.67% in positive subjects versus 33.87% in negative subjects, OR: 3.90 [1.96; 7.79], p=0.0001). Besides, abnormal ALT level is also greater in HCVAb positive subjects than HCVAb negative (66.67%vs.35.27, OR: 3.67 [1.91; 7.05], p=0.0001) (see table 5).

**Table 5.** Liver damage and seroprevalence of hepatitis C antibodies (HCVAb)

|  |  | **HCVAb** |  | **OR [95%CI]** | **p-value** |
| --- | --- | --- | --- | --- | --- |
|  |  | Positive<br>(N=48) | Negative<br>(N=258) |  |  |
|  |  | n (%) | n (%) |  |  |
| **ALT** | Abnormal | **32(66.67)** | 91 (35.27) | 3.67[1.91; 7.05] | 0.0001 |
|  | Normal | 16 (33.3) | 167 (64.73) |  |  |
| **AST** | Abnormal | 31(64.58) | 113(43.8) | 2.34[1.23; 4.44] | 0.009 |
|  | Normal | 17 (35.42) | 145 (56.20) |  |  |
| **ALT+AST** | Abnormal | **30 (62.5)** | 63 (24.4) | 3.90 [1.96; 7.79] | 0.0001 |
|  | Normal | 15 (31.25) | 123 (47.67) |  |  |

## DISCUSSION

In order to investigate the association between HCVAb carriage and liver impairment in an African RLS, a facility-based observational study was conducted from July-August 2021 among individuals attending the “St Monique” Health Center, located at Ottou-village, a rural community of Yaounde, Cameroon. Following a consecutive sampling, consenting individuals were tested for anti-HCV antibodies, hepatitis B surface antigen (HBsAg) and HIV antibodies (HIVAb) as per the national guidelines.

Among the 306 participants surveyed in Ottou-village, at the periphery of Yaounde Cameroon, female were predominant with a percentage of 82.35% (n=252), versus 17.65% (n=54) for males participants [sex ratio (F/M) 5/1]. The mean age was 34.52±3.42 years [min 18, max 72] and subjects aged under 30 years were the most represented (50.98%). Kamga et al in 2018 in the same study site reported a similar female predominance of 68.63% (105/153) and their mean age was 30.4 years ± 5.63 years [26]. While several studies in community settings in Cameroon have also found similar women predominance trend [27, 28, 29, 30], contrary results were more or less reported in rural area of foreign countries including Congo [31], Egypt [32], China [33], in the USA [34]. This is actually the reflect of Cameroonian demographics with in general more women than men and also a relative young population. In addition, it appears as if women seem much more interested in health related matter than men in our context. Furthermore, in this study, more than half (54.9%) of the participants were married and the majority of participants declared having several sexual partners, having experienced a sexually transmitted infection once in their life already and not to regularly use condom. These results underscore the need of continuous sensitization of rural populations on the transmission routes of sexually transmitted diseases.

According to the hepatitis C antibody rapid detection tests results, participants who tested positive were 48 out of 306 (15.69%) [95% CI =11.58% - 19.78%]. HCVAb seroprevalence in Cameroon varies across regions and tribes within the same country. While some studies reported high HCVAb carriage: in the southern 12% [35], in the western6.3% [36], numerous studies on the contrary revealed a relative much lower prevalence: 2.2% anti-HCV in the Northern [37], 0.6% in the eastern[38], 0.4% in the western [39], 4.8% in the Littoral Region [40], 1.44% in the center region [41]. Equally, various burden of anti-HCV antibodies have also been reported in community settings worldwide: 0.77% in Romania [42], 1.02% in Sudan [43], 1.2% in Madagascar [44], 2.4%in Iraq [45], 3.6% in India [46], 19.80% in Cairo [47]. It is known that anti-HCV seropositivity from population-based studies is used to compare levels of HCV infection [9]. According to WHO, countries in Africa and Asia have the highest rate of anti-HCV carriage, whereas industrialized countries in North America, Western Europe, and Australia are known to be lower endemic [48, 49, 50].

The prevalence of liver damage (abnormal ASAT+ALAT) was 30.39% (93/306) with 40.20% (123/306) of abnormal ALAT. Similar studies in rural setting also portrayed high level of serum ALT: 7.1% in Uganda [51], 7.4 %11.2% in Australia [52], 15% in China [53], 22.5% in North Indian [54]. Furthermore, Studies in the USA and Scandinavian revealed about 15% of chronic HCV among liver impaired participants presenting mild to moderate elevations of serum aminotransferase levels for at least 6 months [55, 56]. In fact, serum ALT measurement affords an easily accessible, low cost-effective blood test that is utilized throughout many countries as a liver disease detection tool [21]. ALT is considered as an Indicator of Liver Disease that could globally be used like a valuable screening test for inapparent liver disease, such as asymptomatic viral hepatitis and non-alcoholic fatty liver disease, that still remains largely undiagnosed worldwide [21, 22].

This study portrayed higher prevalence of abnormal level of both ALT + AST as well as abnormal ALT in anti-HCV positive as compared to anti-HCV negative (OR: 3.90 [1.96; 7.79], p=0.0001 and OR: 3.67 [1.91; 7.05], p=0.0001 respectively). Namme et al in 2015 in Douala, Cameroon found 55.1% (246/444) of ALT above upper limit of normal among anti-HCV positive patients [21]. Also, Raheem et al in Nigeria in 2021 found that ALT values were significantly elevated in HCV seropositivity [57]. Equally, Méndez-Navarro et al in Mexico reported 82.6% (289/350) abnormal ALT vs. 17.4% (61/350) normal ALT in 350 consecutive patients with anti-HCV positive between 2003 and 2005 [58]. Many others findings reported higher elevated serum ALT level in anti-AHCV positive individual [59, 60, 61]. In fact, the screening for HCV is routinely strongly recommended in patients with elevated ALT levels and vice-versa [62]. On one side, HCVAb are a commonly available serological marker of HCV infection and on other side, it has been shown that high ALT is a marker for liver disease detection [63]. Chronic hepatitis C is likely associated with variable ALT levels, ranging from normal to high and it is reported that persistently normal levels of ALT in patients with hepatitis C correlates with good prognosis notably lower progression and occurrence of complications like cirrhosis [64]. Furthermore, even though an estimated proportion (25%) of HCV patients have persistently normal ALT levels [65], high ALT *level* remains an excellent tool in predicting viremia in anti-HCV-positive patients after excluding other causes of liver disease [20]. This study prompts the reinforcement of the follow up and management of anti-HCV positive individuals with high ALT level in community setting.

## Conclusion

This study highlights a highly endemic HCVAb rate as well as a concerning burden of liver impairment in this rural health facility. Interestingly, HCVAb carriage is associated with abnormal liver levels of enzyme (ALT/AST), especially among the elderly populations. Hence, in the absence of nuclei acid testing, ALT/AST are relevant sentinel markers to screen HCVAb carriers who require monitoring/care for HCV-associated hepatocellular carcinoma in RLS.

## Data Availability

All data produced in the present study are available upon reasonable request to the authors

## List of abbreviations

ALT: Alanine transaminase
AST: Aspartate Aminotransferase
HBsAg: Hepatitis B Surface Antigen
HCC: Hepatocellular Carcinoma
HCVAb: Hepatitis C Antibody
HCV RT-PCR: Hepatitis C Virus Retrotranscriptase Polymerase Chain Reaction
HIVAb: Human Immunodeficiency Virus Antibody
PLHCV: People Living with Hepatitis C
MEDIBIO-LAB: Medical Biology
RLS: Resource-Limited Setting
WHO: World Health Organization

## Declarations

### Ethics approval and consent to participate

The study was conducted in accordance with the Declaration of Helsinki. The research proposal was evaluated and ethical clearance was obtained from the Regional Ethics Committee for Research involving Humans of the Center Region Cameroon (N°: CE01132/CRERSHC, on May 10^th^, 2021). Additionally, we obtained administrative authorization from the directors of “Sainte Monique” Health Center (Ref N°115/2021/CSGPSM/RS) and MEDIBIO-LAB (N°2021/10/MEDIBIO-LAB/LR) where the study was conducted. An information note was given to all the eligible participants, who then provided their written informed consent before enrollment into the study. The confidentiality of study participants was secured via the use of identification codes.

### Consent for publication

not applicable

### Availability of data and materials

All data are available upon request.

### Competing interests

The authors declare that they have no competing interests

### Funding

This research did not receive any specific grant from funding agencies.

### Authors’ contributions

Designed the study: RKW, GPT, LAKS, EPTL, DB, JF; Planned and performed the experiments: RKW, GPT, DPDD, GFMT, DB; Analysed and interpreted the data: GPT, DPDD, CIYW; Initiated the manuscript: RKW, GPT, LAKS, DPDD, CIYW, DB, JF; Revised the manuscript: All the authors; Approved the final version of the manuscript: All the authors

## Acknowledgements

We hereby would like to thank all participants, Health personnels of “Sainte Monique-Ngonmeda” Health Center and MEDIBIO-Laboratory.

## Authors’ information (optional)

not applicable

1. Department of Microbiology and Parasitology, Faculty of Science, University of Buea, Buea, Cameroon
2. American Society for Microbiology (ASM), ASM Cameroon, Bangangte, Cameroon;
3. Ecole de Santé Publique, Université Libre de Bruxelles, Bruxelles, Belgique ;
4. Department of Microbiology, Faculty of Health Science, University of Buea, Buea, Cameroon;
5. Faculty of Medicine, Sorbonne University, Paris, France;
6. Adventist Cosendai University, Yaounde, Cameroon;
7. Laboratory of Fundamental Virology, Centre for Research on Emerging and Reemerging Diseases (CREMER), Yaounde, Cameroon;
8. Yaounde Central Hospital, Yaounde, Cameroon ;
9. Faculty of Health Sciences, University of Buea, Buea, Cameroon;
10. Virology Laboratory, Chantal BIYA International Reference Centre for research on HIV/AIDS prevention and management (CIRCB), Yaoundé, Cameroon;
11. Faculty of Medicine and Biomedical Sciences, University of Yaounde I, Yaounde, Cameroon.

## Notes

### Competing Interest Statement

The authors have declared no competing interest.

### Funding Statement

This study did not receive any funding

### Author Declarations

Ethics committee/IRB of the Center Region Cameroon gave ethical approval for this work

